# Antibody response to SARS-CoV-2 vaccination is extremely vivacious in subjects with previous SARS-CoV-2 infection

**DOI:** 10.1101/2021.03.09.21253203

**Authors:** Annapaola Callegaro, Daniela Borleri, Claudio Farina, Gavino Napolitano, Daniela Valenti, Marco Rizzi, Franco Maggiolo

## Abstract

**Background:** The SARS-CoV-2 pandemic calls for rapid actions, now principally oriented to a world-wide vaccination campaign.

In this study we verified if, in individuals with a previous SARS-CoV-2 infection, a single dose of mRNA vaccine would be immunologically equivalent to a full vaccine schedule in naïve individuals.

**Methods:** Health care workers (184) with a previous SARS-CoV-2 infection were sampled soon before the second dose of vaccine and between 7 and 10 days after the second dose, the last sampling time was applied to SARS-CoV-2 naïve individuals, too.

Antibodies against SARS-CoV-2 were measured using Elecsys^®^ Anti-SARS-CoV-2 S immunoassay.

The study was powered for non-inferiority. We used non parametric tests and Pearson correlation test to perform inferential analysis.

**Results:** After a single vaccine injection, the median titer of specific antibodies in individuals with previous COVID-19 was 30,527 U/ml (IQR 19,992-39,288) and in subjects with previous SARS-CoV-2 asymptomatic infection was 19,367.5 U/ml (IQR 14,688-31,353) (P=0.032). Both results were far above the median titer in naïve individuals after a full vaccination schedule: 1,974.5 U/ml (IQR 895-3,455) (P<0.0001). Adverse events after vaccine injection were more frequent after the second dose of vaccine (mean 0.95, 95%CI from 0.75 to 1.14 versus mean 1.91, 95%CI from 1.63 to 2.19)(P<0.0001) and in exposed compared to naïve (mean 1.63; 95%CI from 1.28 to 1.98 versus mean 2.35; 95%CI from 1.87 to 2.82)(P=0.015).

**Conclusion:** In SARS-CoV-2 naturally infected individuals a single mRNA vaccine dose seems sufficient to reach immunity. Modifying current dosing schedules would speed-up vaccination campaigns.

## Introduction

Severe acute respiratory syndrome coronavirus 2 (SARS-CoV-2), which was identified in China in December 2019, causes coronavirus disease 2019 (COVID-19), a severe, acute respiratory syndrome with a complex, highly variable disease pathology. The severe and worldwide effect of the pandemic on human society calls for rapid actions, now principally oriented to a world-wide vaccination campaign.

Two SARS-CoV-2 spike mRNA vaccines received emergency use authorization by the FDA in December 2020 (BNT162b2/Pfizer; mRNA-1273/Moderna) [2]. Both Phase 3 trials on these vaccines reported high efficacy in preventing symptomatic SARS-CoV-2 infections after two doses of the vaccine administered three to four weeks apart in subjects without previous SARS-CoV-2 infection [3,4]. However, little is known about subjects with previous exposure to SARS-CoV-2. Anecdotally, individuals with pre-existing immunity experience more severe reactogenicity after the first doses compared to naïve individuals.

The present study was designed to verify if, in individuals with a previous SARS-CoV-2 infection, a single administration of BNT162b2/Pfizer vaccine would elicit an immunological response superimposable to a full vaccine schedule in naïve individuals, assuming that, for individuals with pre-existing immunity to SARS-CoV-2, the first vaccine dose could immunologically resemble the booster dose in naïve individuals.

## Methods

The study was carried out on 184 health care workers. Individuals with a previous COVID-19 symptomatic disease or a SARS-CoV-2 asymptomatic infection were sampled for measuring SARS-CoV-2 antibody responses soon before the second dose of vaccine and between 7 and 10 days after the second dose, the last sampling time was applied to SARS-CoV-2 naïve individuals, too. Antibodies against SARS-CoV-2 in serum samples were measured using Elecsys^®^ Anti-SARS-CoV-2 S immunoassay (Roche Diagnostics GmbH, Sandhofer Strasse 116, D-68305 Mannheim) for the in vitro quantitative determination of antibodies (including IgG) to the SARS-CoV-2 spike (S) protein receptor binding domain (RBD) [5,6], according to the manufacturer’s instructions. Briefly, all serum samples were determined with ≥ 0.8 U/ml by the Elecsys^®^ Anti-SARS-CoV-2 S test and the values above the measurement range (0.40 >250 U/mL) were quantified using the recommended “Diluent Universal” on Cobas e602 analyzer. As reported by Roche Diagnostics, the specific U/mL units of the Roche Elecsys® Anti-SARS-CoV-2 S test can be considered equivalent to the binding antibody units (BAU)/mL of the first WHO International Standard for anti-SARS-CoV-2 immunoglobulins [7,8].

### Statistical analysis

The study was powered to demonstrate no-inferiority of a single dose in pre-exposed subjects compared to two injections in naïve individuals, provided that the magnitude of immunological response was equivalent in the 2 groups. Data are summarized as medians and IQR, means and 95% confidence intervals or percentages. We used Mann-Whitney test, Wilcoxon rank test and Pearson correlation test to perform inferential analysis.

### Ethics

The Provincial Ethical Committee approved the study and all participants gave their informed consent.

### Founding

The study has no funding

## Results

Out of 184 health care workers, joining the National vaccination campaign, 53 were previously diagnosed with COVID-19; 21 had been previously tested positive (nasal swab or serological test) for SARS-CoV-2 without any symptom of COVID-19 and 110 were naïve individuals. Overall, 125 (67.9%) were of feminine gender and the median age was 50 years (IQR 39-56). All received two doses of BNT162b2/Pfizer vaccine. After a single vaccine injection, the median titer of specific antibodies in individuals with previous COVID-19 was 30,527 U/ml (IQR 19,992-39,288) and in subjects with previous SARS-CoV-2 asymptomatic infection was 19,367.5 U/ml (IQR 14,688-31,353) (P = 0.032). Both results were far above the median titer in naïve individuals after a full vaccination schedule: 1,974.5 U/ml (IQR 895-3,455) (P < 0.0001)(figure 1). With the second dose, in previously exposed individuals, median titers raised to 43,073 U/ml (IQR 31,605-61,903) (P < 0.0001). Considering the correlation between the specific U/mL units of the Roche Elecsys® Anti-SARS-CoV-2 S test and the BAU/mI, a regression analysis confirmed a strict correlation up to 1000 BAU/ml, the value assigned to WHO International Standard. As all but two individuals with previous SARS-CoV-2 infection and most naïve individuals showed antibody titers higher than 1000 BAU/ml, we used throughout the study the definition U/ml.

**Figure 1:**
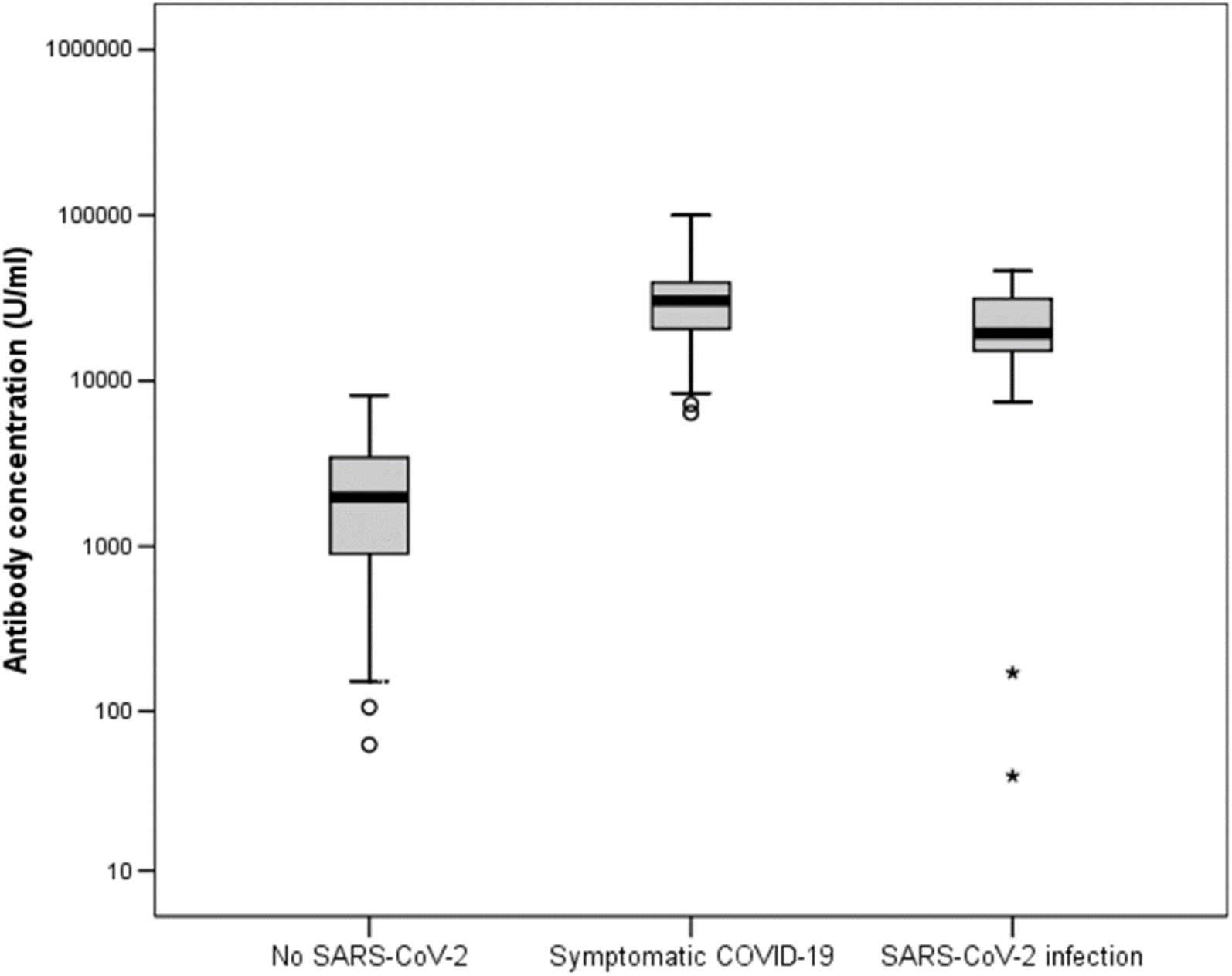
Specific antibody titers after a full vaccination schedule (2 injections) in SARS-CoV-2 naïve individuals and a single vaccine injection in subjects with previous exposure to SARS-CoV-2 either symptomatic or not

Titers were slightly higher in subjects who acquired the infection during the first wave, observed in March/April 2020, 8-11 months before testing, compared to those with more recent infection (2-3 months) (P = 0.017), but titers were not influenced by age (P = 0.083). Adverse events after vaccine injection were more frequent after the second dose of vaccine (mean 0.95, 95%CI from 0.75 to 1.14 versus mean 1.91, 95%CI from 1.63 to 2.19)(P < 0.0001). Adverse events were also more frequent in exposed compared to naïve individuals both after the first (mean 0.77; 95%CI from 0.55 to 1.00 versus mean 1.23, 95%CI from 0.89 to 1.50)(P = 0.002); or the second (mean 1.63; 95%CI from 1.28 to 1.98 versus mean 2.35; 95%CI from 1.87 to 2.82) (P = 0.015) vaccine dose (figure 2).

**Figure 2:**
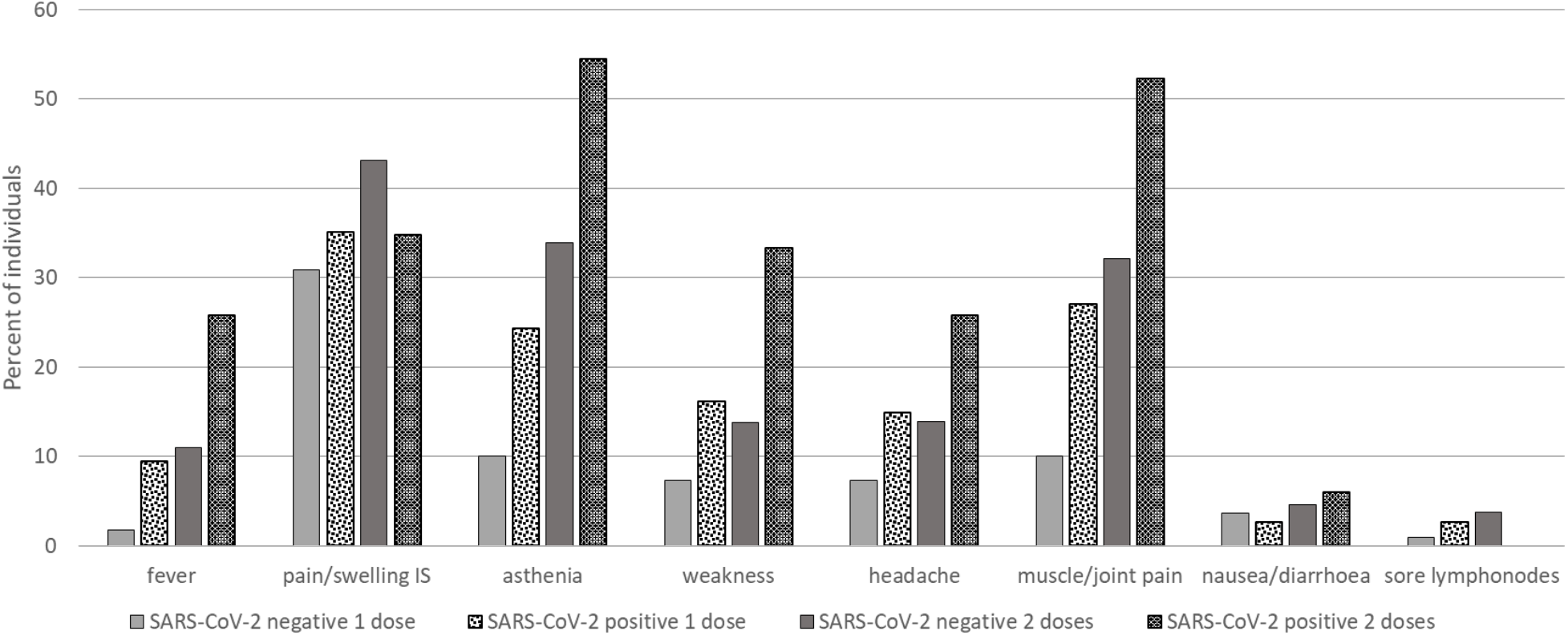
Most frequent AEs observed after vaccine injection

## Discussion

A world-wide vaccination campaign is an expensive, time and labor consuming effort. Eventually, considering a single vaccine dose in subjects previously exposed to SARS-CoV-2 would offer advantages in terms of costs, timing and possibility to reach a greater number of subjects in a shorter period of time.

Our findings suggest that a single dose of mRNA vaccine elicits a very strong immune response in seropositive individuals with post-dose antibody titers 10-fold higher to those observed in naïve individuals who received a full vaccination schedule. Interestingly, this type of response is present whether or not individuals developed a symptomatic COVID-19 disease. These observations are in line with the hypothesis that the first vaccine dose serves as a boost in naturally infected individuals [9]. Our study has some limitations. Being conducted in health care workers, the age of enrolled individuals ranged from 24 to 66 years and we cannot exclude that younger or older subjects could react differently. Furthermore, most of our subjects were Caucasian and we therefore cannot extend our observation to different ethnicities. Finally, we did not observe any non-responder to the vaccine, although this possibility cannot be absolutely excluded. However, in our experience all patients but two with a pre-exposure to SARS-CoV-2 showed antibody titers well above the 1000 IU/ml after a single vaccine dose. That appears reassuring in the light of a differentiated vaccination schedule.

In SARS-CoV-2 naturally infected individuals a single mRNA vaccine dose seems sufficient to reach immunity. Modifying currently implemented dosing schedules would speed-up vaccination campaigns and would limit occurrence of adverse events in this sub-group of individuals

## Data Availability

All data are available in an electronic database on request to the principal investigator

